# Machine learning methods applied to genotyping data capture interactions between single nucleotide variants in late onset Alzheimer’s disease

**DOI:** 10.1101/2021.08.30.21262815

**Authors:** Magdalena Arnal Segura, Dietmar Fernandez Orth, Claudia Giambartolomei, Giorgio Bini, Eleftherios Samaras, Maya Kassis, Fotis Aisopos, Jordi Rambla De Argila, Georgios Paliouras, Peter Garrard, Gian Gaetano Tartaglia

## Abstract

**INTRODUCTION:** Genome-wide association studies (GWAS) in late onset Alzheimer’s disease (LOAD) provide lists of individual genetic determinants. However, GWAS are not good at capturing the synergistic effects among multiple genetic variants and lack good specificity.

**METHODS:** We applied tree-based machine learning algorithms (MLs) to discriminate LOAD (> 700 individuals) and age-matched unaffected subjects using single nucleotide variants (SNVs) from AD studies, obtaining specific genomic profiles with the prioritized SNVs.

**RESULTS:** The MLs prioritized a set of SNVs located in close proximity genes PVRL2, TOMM40, APOE and APOC1. The captured genomic profiles in this region showed a clear interaction between rs405509 and rs1160985. Additionally, rs405509 located in APOE promoter interacts with rs429358 among others, seemingly neutralizing their predisposing effect. Interactions are characterized by their association with specific comorbidities and the presence of eQTL and sQTLs.

**DISCUSSION:** Our approach efficiently discriminates LOAD from controls, capturing genomic profiles defined by interactions among SNVs in a hot-spot region.

## Background

Alzheimer’s disease (AD) is a neurodegenerative pathology and the most common cause of late life dementia with symptoms such as memory loss, language deficits, disorientation, mood changes, and in advanced stages is associated with loss of vegetative function and, eventually, death^1^. Approximately the 5% of the total number diagnosed with AD develop symptoms of dementia between the ages 45 and 65, and are designated as early onset Alzheimer’s disease (EOAD)^2^. Conversely, the prevalence of the disease in the population aged above 65 currently represent around 95% of the total AD cases, and are designated as late onset Alzheimer disease (LOAD)^3^. AD is found in about 1 in 8 people aged 65 to 74, and the number doubles every five years after age 65, reaching 1 in 2 people over 85 y.o ^1^. At the pathophysiological level, AD is defined by plaque accumulation of anomalous folded amyloid beta protein outside neurons, and abnormal aggregation of the tau protein inside cells^4^. These two events induce the loss of neurons and synapses in the cerebral cortex and certain subcortical regions, resulting in the cognitive impairments perceived in AD patients.

The genomic characterization of AD has improved in the last decades thanks to the emergence of genome-wide association studies (GWAS)^5^. However, these tools miss the synergistic effects caused by various genomic loci and lack good specificity due to the multiple testing problem and linkage disequilibrium (LD) issue^6^. In this context, the selection of genetic determinants for the follow-up in laboratory and clinical studies remains a challenge, and most of the mechanisms in which the discovered predisposing and protective genetic alterations contribute to AD, are still unknown^7^. In the case of LOAD, heritability is estimated to be around 56-79%^8^, and APOE polymorphic alleles are the major genetic determinants of susceptibility discovered until now^9 10^. Nevertheless, there are more candidate genes such as TOMM40, PVRL2, ABCA7, ADAM10, BIN1, CLU and CR1 among others^5 11^, and nowadays, there is a growing consensus considering LOAD a polygenic risk disease^12^.

Machine learning (ML) methods are growing in popularity for their contributions to a wide range of fields including medicine ^13 14^. ML classifiers have been previously implemented to classify AD using genotyping data reaching an accuracy of 0.84^15 16^. Additionally, they have been used in the post-GWAS prioritization of genomic variants in several diseases^17 18 19^. As ML algorithms work better with a limited set of predictors in order to be efficient, and the full set of single nucleotide variants (SNVs) in genotyping arrays is too large to reach a reasonable computational performance, a set of AD related SNVs are typically pre-selected and used as predictors in the ML models. As input variables, ML approaches can accept a list of SNVs without any prior assumptions about the genetic contribution to the traits and the method itself calculates the importance of the SNVs during the learning step.

In this study, our initial aim was to classify individuals with LOAD and controls without any neurodegenerative disease both from UKB^20^, using ML methods and data from genotyping arrays. Our second aim was to select the SNVs with higher feature importance and retrieve a set of genomic profiles that are related to AD. We did a first selection of genomic variants considering previously reported SNVs related to AD in the DisGeNet^21^ database. DisGeNet integrates data from curated resources such as ClinVar^22^, the GWAS Catalog^23^ and GWASdb^24^. As for the classification method, we tested three tree-based ML approaches, Gradient Boosted Decision Trees (GB), Extremely Randomized Trees (ET) and Random Forest (RF). Tree based algorithms perform yes/no decisions in branches leading to a sample’s classification, which is particularly appropriate with categorical predictors such as SNVs. Here we show the utility of tree-based ML methods to classify LOAD, to prioritize a small set of SNVs related to the disease, and to draw distinct LOAD genomic profiles based on the interactions between these SNVs.

## Methods

### Sample selection and clinical information in UKBiobank

From UKB^20^, a total number of 738 participants with AD and y.o > 70, were selected using ICD-10 codes representative of AD excluding the EOAD ICD-10 codes from the available hospitalization records (**Supplementary Table 1, page 1)**. In addition, 75000 participants were selected as controls > 70 y.o and without any reported mental and behavioral disorder (ICD-10: F00–F99) or disease of the nervous system (ICD-10: G00–G99) in hospitalization records. Participants that requested to withdraw from UKB were excluded. Gender and sex distribution of selected samples across conditions is shown in **Supplementary Figure 1 a and b**. Comorbidities were retrieved for the selected individuals (LOAD and controls) excluding the ICD-10 categories F00–F99 and G00–G99.

### Pre-processing and selection of genomic variants

Genome-wide genotyping data from “Affymetrix UK BiLEVE Axiom array” and “Affymetrix UK Biobank Axiom® array” (BED, BIM and FAM files) available for the 500.000 participants in the UK Biobank cohort was used as the source of genomic data. The pre-processed genotyping data (zygosity, haplotype estimation and genotype imputation) was available to download from UKB. Bed, Bim and Fam files annotated with annovar^25^ were used to extract individualized genotyping data. In terms of quality assessment, genomic variants with MAF ≤ 0.01% and Hardy-Weinberg Equilibrium (HWE) p-value < 10E-4 were filtered out. Monomorphic markers (SNVs with the same genotype in all subjects) were also excluded. The list of SNVs to be used as predictors was obtained from the “curated variant disease associations” dataset in DisGeNet^21^, filtering for the AD categories described in **Supplementary Table 1, page 2**. A total number of 145 SNVs reported to be related to AD and passing the quality filters mentioned above were selected as AD predictors. The annotated list of AD related predictors, is provided in **Supplementary Table 2** and the distribution over chromosomes and genomic regions is shown in **Supplementary Figure 1 c and d**. After the selection of predictors, numeric matrices were built with rows representing samples and columns SNVs. We used the dbSNP ID as unique identifier for SNVs. In the matrix, SNVs were categorized as 1, 2 or 3 corresponding to the three possible genotypes, minor allele of the SNV absent, present in one allele or present in both alleles respectively. Missing values were categorized with 0.

### ML models: building and evaluation

Python 3.7.6 with Scikit-learn v0.22.1 module was used to build the ML models on the pre-processed matrix described in the section “Pre-processing and selection of genomic variants”. A train/test split was applied to have 80% of samples for training and 20% of samples for testing. Samples were balanced to have the same proportion of LOAD and controls using random undersampling. Hyper-parameter selection was performed on the training set through a 10-fold cross-validation. Two metrics such as AUC-ROC and f-score were considered for determining the best hyper-parameter configuration for each model. The median of AUC-ROC (**Supplementary Figure 2 a, b, c**) and f-score (**Supplementary Figure 2 d, e, f**) was higher than 0.7 in validation sets of models with AD predictors. The final model with the optimized parameters was trained on the original train set (80%) and tested on the test set (20% of samples). The selected hyper-parameter values are listed in **Supplementary Table 1, page 3**.

### Statistical test for interactions between pairs of SNVs

For all the possible pairwise combinations of 6 SNVs, R (v4.0.4) was used to build full generalized lineal models (glm) considering two SNVs as independent variables with their individual effect and interaction to classify LOAD and controls, and a reduced glm with the same variables but considering only the individual effect of each SNV without the interaction term to classify both classes. The models were built with samples that didn’t have any missing value in any of the 6 SNVs, consisting in a total number of 622 LOAD samples and 622 randomly selected controls. The function anova.glm in stats package was used to perform the analysis of deviance between the full glm and reduced glm, comparing the reduction in deviance with a chi-squared test.

## Results

### Evaluation metrics across different ML models

We tested the ability of GB, ET and RF models with the set of SNVs defined as AD predictors to classify LOAD and controls. The evaluation was made with 7 different metrics shown in **Table 1**. The best scores in all evaluation metrics were obtained using GB with an accuracy, f-score, sensitivity, specificity, positive predicted value (PPV) and negative predicted value (NPV) of 0.80, and an AUC-ROC of 0.91. RF performed slightly better than ET in accuracy (0.74), f score (0.74), sensitivity (0.73), specificity (0.75), PPV (0.75), and NPV (0.73), but AUC-ROC was better in ET (0.82) (**Supplementary Figure 3 a)**.

**Table 1.** Summary of the evaluation metrics obtained with GB, ET and RF models and AD predictors. PPV = Positive Predicted Value, NPV = Negative Predicted Value. ML models with best scores in each evaluation metric are highlighted in red.

|  | Accuracy | AUC-ROC | F score | Sensitivity | Specificity | PPV | NPV |
| --- | --- | --- | --- | --- | --- | --- | --- |
| **GB** | **0.801** | **0.912** | **0.800** | **0.797** | **0.804** | **0.803** | **0.799** |
| **ET** | 0.707 | 0.820 | 0.706 | 0.703 | 0.710 | 0.708 | 0.705 |
| **RF** | 0.739 | 0.804 | 0.735 | 0.725 | 0.754 | 0.746 | 0.732 |

### Prioritization of SNVs using feature importance

To identify the SNVs that provide the strongest signal for the classification, we ranked the AD predictors based on the impurity-based feature importance (FI). For each ML method, a FI higher than 0.01 was used to select the SNVs among the 145 in AD predictors considered as relevant during the classification (**Supplementary Figure 3 b to d**). A set of 9, 20 and 15 SNVs were selected in GB, ET and RF models respectively (**Supplementary Table 3**). We named the SNVs using their dbSNP ID and referring to the presence or absence of the minor allele. Overall, all the prioritized SNVs except one, rs7561528, prioritized by RF in chromosome 2, were located in a region of chromosome 19 comprising PVRL2, TOMM40, APOE and APOC1 genes (**Figure 1 a**). The intersection of the prioritized SNVs between the three ML methods is shown in **Figure 1 b**. The 6 SNVs prioritized by the three ML models were: rs1160985 in TOMM40; rs405509, rs7412, rs769449 and rs429358 in APOE, and rs4420638 downstream APOC1 (**Table 2**).

**Figure 1.**
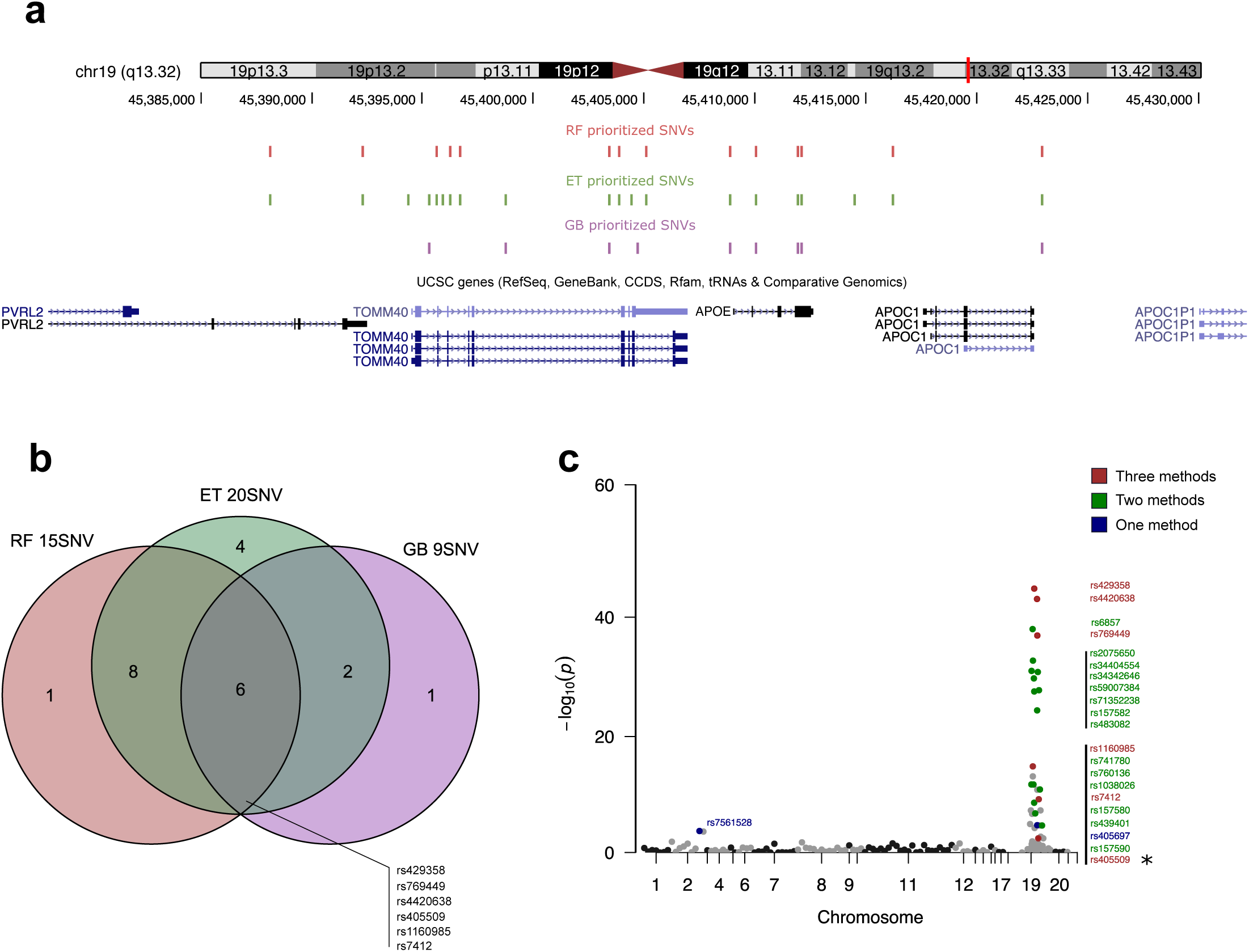
**a** The genomic location of SNVs selected using a FI > 0.01 in the chromosome19 hot-spot region. SNVs prioritized by different ML methods are illustrated in different tracks. **b** Venn diagram showing the intersection of the prioritized SNVs by GB, ET and RF. The name of the SNVs in the intersection with the three methods is provided. **c** For the 145 SNVs in AD predictors, distribution of the Fisher-test p-values obtained measuring differences in AF between LOAD and controls over the chromosomes. The name of the SNVs prioritized by any of the three ML methods is provided and a color is assigned depending on the number of times a SNV was selected by any one of the methods. The 6 SNVs prioritized by GB, ET and RF are colored in red.

**Table 2.**
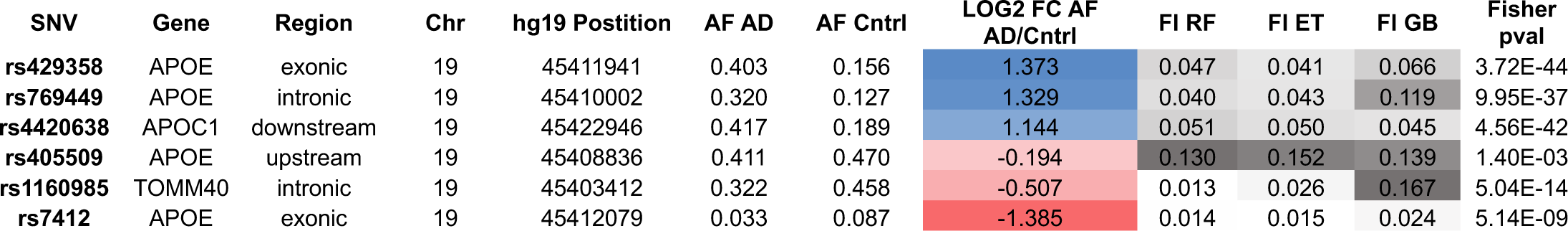
Characteristics of the list of 6 SNVs prioritized by the three ML methods. dbSNP ID together with gene annotations are provided in the columns “SNV”, “Gene”, “Region”, “Chr” and “hg19 Position”. AF in AD and in controls are used to calculate the log2FC of AF in AD vs Cntrl (column “LOG FC AF AD/Cntrl”). SNVs are ordered from the highest logFC (top) to the lowest (bottom) and colored in blue and red accordingly. FI obtained in RF, ET and GB are in columns “FI RF”, “FI ET” and “FI GB” respectively. Fisher test p-values measuring the significance of AF differences between AD and controls are provided in the “Fisher pval” column.

| SNV | Gene | Region | Chr | hg19 Postition | AF AD | AF Cntrl | LOG2 FC AF AD/Cntrl | FI RF | FI ET | FI GB | Fisher pval |
| --- | --- | --- | --- | --- | --- | --- | --- | --- | --- | --- | --- |
| rs429358 | APOE | exonic | 19 | 45411941 | 0.403 | 0.156 | 1.373 | 0.047 | 0.041 | 0.066 | 3.72E-44 |
| rs769449 | APOE | intronic | 19 | 45410002 | 0.320 | 0.127 | 1.329 | 0.040 | 0.043 | 0.119 | 9.95E-37 |
| rs4420638 | APOC1 | downstream | 19 | 45422946 | 0.417 | 0.189 | 1.144 | 0.051 | 0.050 | 0.045 | 4.56E-42 |
| rs405509 | APOE | upstream | 19 | 45408836 | 0.411 | 0.470 | -0.194 | 0.130 | 0.152 | 0.139 | 1.40E-03 |
| rs1160985 | TOMM40 | intronic | 19 | 45403412 | 0.322 | 0.458 | -0.507 | 0.013 | 0.026 | 0.167 | 5.04E-14 |
| rs7412 | APOE | exonic | 19 | 45412079 | 0.033 | 0.087 | -1.385 | 0.014 | 0.015 | 0.024 | 5.14E-09 |

We used the Fisher-test p-value to measure the differences in allele frequency (AF) between LOAD and controls and then check if the SNVs with higher FI were also the ones with higher AF differences. Among all AD predictors, the highest AF differences were in a set of SNVs located in chromosome 19 (**Figure 1 c)**, in a hot-spot region comprising PVRL2, TOMM40, APOE and APOC1 genes, the same region where the prioritized SNVs using FI were located (**Figure 1 a**). When looking at the AF of SNVs prioritized by the three methods in **Table 2**, SNVs more frequent in LOAD with respect controls were rs429358 (pval=3.72E-44 logFC=1.37), rs769449 (pval=9.95E-37 logFC=1.33) and rs4420638 (pval=4.56E-42 logFC=1.14). Conversely, the SNVs more frequent in controls were rs405509 (pval=1.40E-03 logFC=-0.19), rs1160985 (pval=5.04E-14 logFC=-0.51), and rs7412 (pval=5.14E-09 logFC=-1.39). rs429358 had the highest AF difference between conditions (**Figure 1 c**), being 2.6 times more frequent in LOAD with respect to controls (logFC=1.37). Yet, rs429358 was not the SNV with the highest FI in any of the ML methods (**Table 2** and **Supplementary Table 3)**. Alternatively, rs405509 reached the highest FI in RF (0.13) and ET (0.15) and the second higher in GB (0.14), but had the lowest AF differences between LOAD and controls compared with the other prioritized SNVs (logFC=-0.194) (**Figure 1 c** and **Table 2)**. We hypothesized the importance of rs405509 in the ML classification is probably due to the co-occurrence or mutual exclusion with other variants that together form certain genomic profiles, rather than for being more present in one condition with respect to the other.

### Interactions in the hot-spot region of chromosome 19

To identify possible interactions occurring between the sets of prioritized SNVs, we analyzed the genomic profiles whose samples were correctly classified as true positives (TP) or true negatives (TN) all the times in GB, ET and RF (**Figure 2, Supplementary Figure 4** and **Supplementary Figure 5** respectively). Most of the patterns described hereafter are observed in genomic profiles captured by the three ML methods. However, for simplicity we discuss the genomic profiles defined by GB only **Figure 2**. This decision is supported by the fact that GB was the model with best performance in the classification.

**Figure 2.**
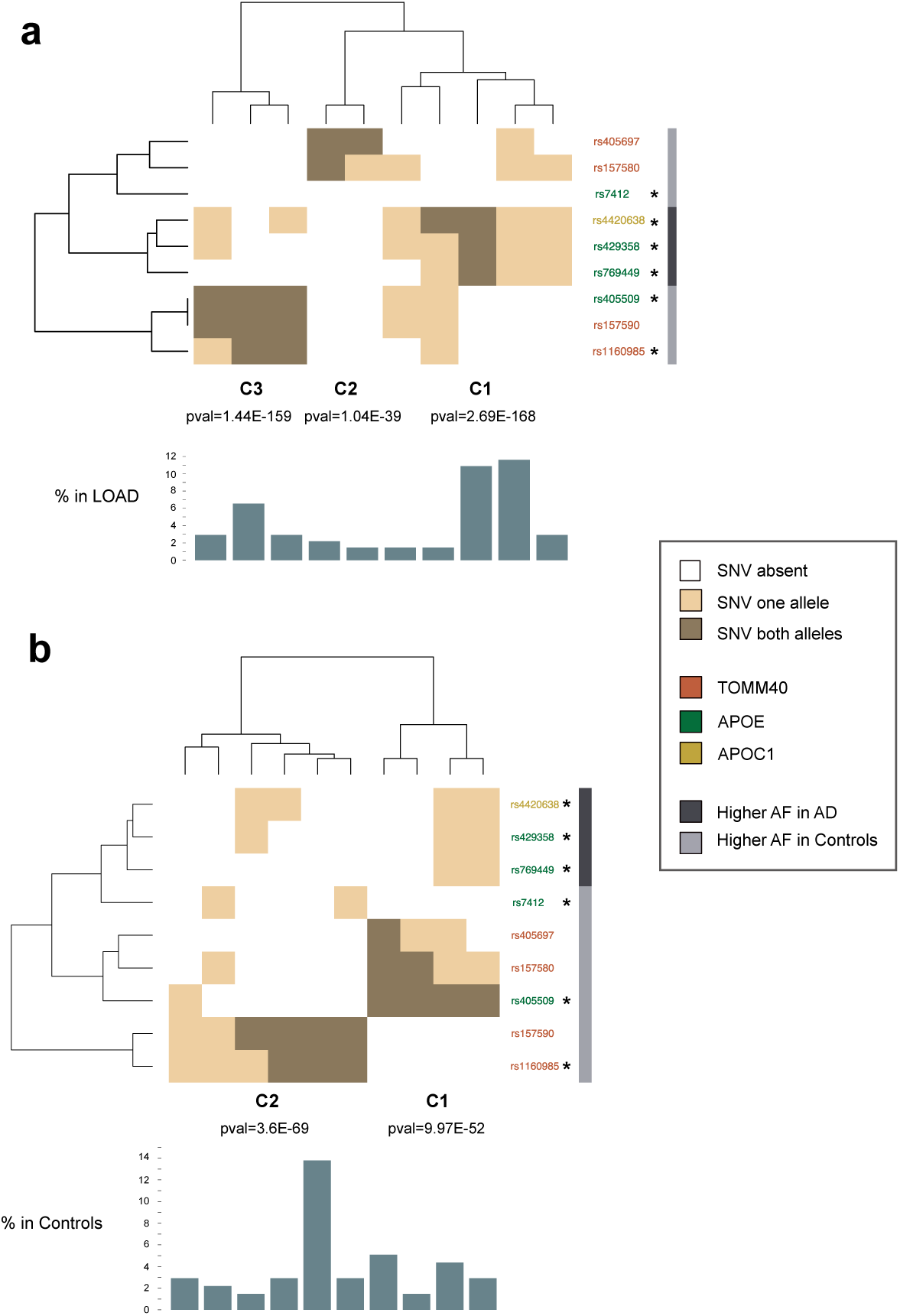
Genomic profiles of correctly classified samples in GB defined with the 9 prioritized SNVs. Genomic profiles with only one sample or having missing values were excluded. In **a**, genomic profiles of true positives (TP) represent all samples that were correctly classified as LOAD. In **b**, genomic profiles of true negatives (TN) represent all samples that were correctly classified as controls. Dendrograms on the top and the left were made with Ward-D2 method and Euclidean distances. Clusters of genomic profiles are indicated with numbers in the X-axis. Fisher-test p-values are provided measuring the statistical significance of different representation of AD and controls in clusters of genomic profiles. The % of samples having each genomic profile in LOAD and controls is indicated in the bar-plots below the heatmaps. SNVs are colored with their corresponding gene loci and information of the higher AF in LOAD or controls is provided in the right-side bar. An asterisk points to the 6 SNVs commonly prioritized by GB, ET and RF.

The majority of TP profiles were characterized as having the three SNVs, rs429358, rs4420638 and rs769449 either in one or two alleles (**Figure 2a C1)**. These genomic profiles were present in the 27.83% of LOAD and 1.21% of controls in the full UKB dataset. The genomic profiles with rs405509 in both alleles co-occurring with rs1160985 were present in the 12.62% of LOAD and were not present in controls (**Figure 2a C3**). Interestingly, rs405509 in both alleles and rs1160985 were mutually exclusive in TN (**Figure 2b C1 and C2**). Moreover, the predisposition to AD caused by the presence of rs429358, rs4420638 and rs769449 in one allele was neutralized with the presence of rs405509 in both alleles in a group of TN (**Figure 2b C1)**. Only GB captured profiles of TP characterized with rs405697 and rs157580 co-occurring either in one or two alleles without the presence of the other 7 SNVs in 3.24% of LOAD (**Figure 2a C2)**.

Altogether, the LOAD genomic profiles captured by ML models suggested that an interaction may exist between the SNVs in the chromosome19 hot-spot region. In order to test the significance of the interactions, we built generalized linear models (glm) to classify LOAD and controls with and without the interaction term using pairwise combinations of SNVs, and we measured the changes in the deviance between both models. We evaluated the interactions on the 6 SNVs prioritized by the three ML methods, on the grounds that these are predictors prioritized by three independent methods and therefore have the strongest signal in the classification. Also, they define the major genomic profiles in our data (**Table 3)**. In accordance with what it was observed in the genomic profiles of TP and TN, the most significant interaction was observed between rs1160985 and rs405509 (pval=1.42E-153) followed by the interactions between rs405509 and the other SNVs located in APOE, rs769449 (pval=1.50E-65), rs429358 (pval=4.39E-21) and rs7412 (pval=3.59E-20).

**Table 3.**
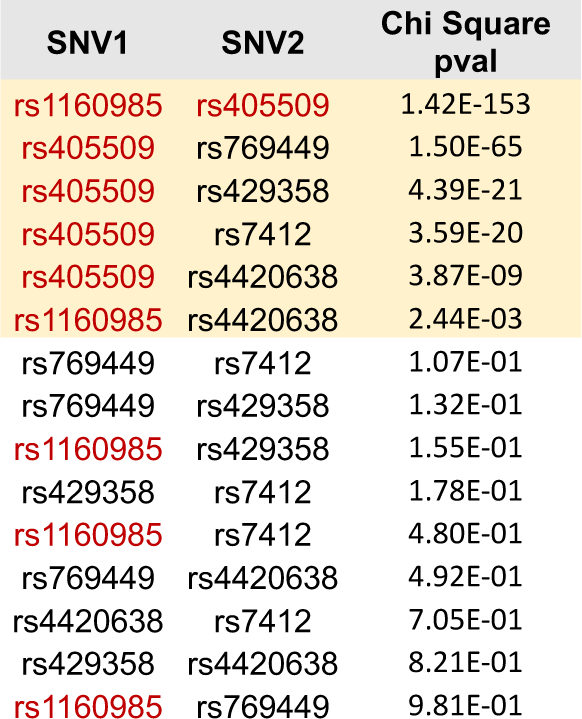
Pairwise test of interactions for the 6 SNVs commonly prioritized by ML models (s1160985 in TOMM40; rs405509, rs7412, rs769449 and rs429358 in APOE, and rs4420638 downstream APOC1). Comparisons were made between the full glm built with two SNVs considering the individual effect and interactions to predict AD and controls, and the reduced glm only considering the individual effect of the two SNVs to predict both conditions. The differences between the full and reduced glm were measured with an anova and the corresponding chi-squared test pval is provided for each comparison. We highlight in red the two SNVs (rs1160985 and rs405509) with 1) high FI scores in the ML models, 2) low AF differences between AD and controls, 3) involved in interaction patterns of genomic profiles.

### Comorbidities present in LOAD with different interaction patterns

We identified two major interaction patterns in LOAD. Interaction 1 (**Figure 2a C3**) characterized by the presence of rs405509 and rs1160985 (in both alleles), and interaction 2 (**Figure 2a C1)** characterized by the absence of rs405509 (in both alleles) and rs7412, as well as the presence of rs769449, rs429358 or rs4420638. LOAD individuals with interaction 1 showed more cases of arthritis and gastritis compared with other LOAD or controls, which may indicate a tendency towards inflammation. By contrast, LOAD individuals with interaction 2 were associated with cases of abnormal weight loss, acute lower respiratory infection, hypothyroidism and atherosclerotic heart disease compared with controls. Conditions such as hypertension, a personal history of diseases of the circulatory system, pure hypercholesterolaemia, anemia and personal history of psychoactive substance abuse were enriched in both LOAD groups compared with controls (**Figure 3** and **Supplementary Table 4**).

**Figure 3.**
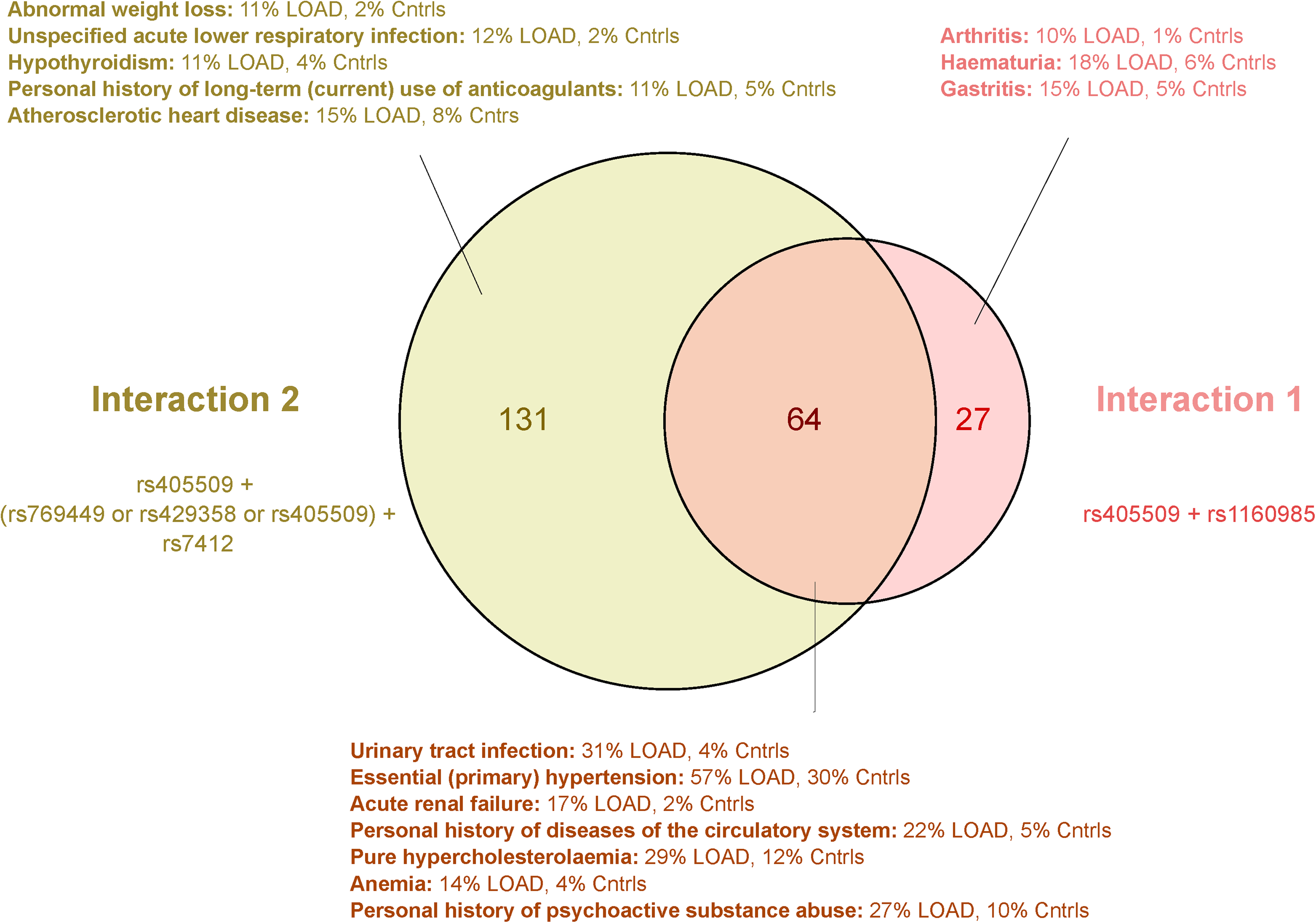
Venn diagram showing diseases and conditions enriched in LOAD individuals having two distinct interaction patterns (interaction 1 and interaction 2). A cut-off pval < 0.001 in the Fisher test was used to select the categories more represented in LOAD in comparison with controls. Subgroups are: LOAD with interaction 1 in light red, LOAD with interaction 2 in green, intersection of both LOAD in dark red. The most representative diseases and conditions present in more than 10% of individuals in the LOAD subgroups are shown with their corresponding percentages.

### eQTL and sQTL in the prioritized SNVs

We examined the effect of the 6 SNVs on gene expression (eQTL) and splicing (sQTL) on different tissues in GTEx^26^ with a pval<1E-4. In terms of gene expression, rs1160985, rs405509, rs7412, rs4420638 were eQTLs of APOE in skin tissue and in the case of rs1160985 heart tissue as well (**Supplementary Table 1, page 5)**. rs429358, rs769449 and rs4420638 were eQTLs of the upstream gene APOC1 in Esophagus, Adrenal Gland and Skin. Even if there were no eQTLs captured in brain tissue for any of the 6 SNVs, this data evidence the presence of a transcriptional regulatory hub in the hot-spot region of chromosome 19 that may be altered by the presence of alternative alleles in the prioritized SNVs. On the other side, all the SNVs except for rs7412 were sQTLs to TOMM40 in brain (**Supplementary Table 1, page 6)**. In addition, rs405509 and rs429358 were sQTL to APOE in lung and brain respectively.

## Discussion

Using tree-based ML methods and the set of 145 SNVs related to AD reported in databases, we were able to classify LOAD and controls, reaching an accuracy of 0.80 and an AUC-ROC of 0.91 in GB. We prioritized a set of 9, 20 and 15 SNVs in GB, ET and RF respectively, from which 6 SNVs were commonly prioritized across the three methods. The 6 SNVs were located in a chromosome19 hot-spot region comprising TOMM40, APOE and APOC1 genes. rs429358 is the most well-characterized LOAD genetic determinant^27 10^, rs7412 is known to be protective against AD^28^, and the two SNVs define the distinct apolipoprotein E (ApoE) isoforms^29^. rs4420638 is in strong LD with rs429358^9^ and for this reason, its link with AD is attributed to rs429358. rs769449 has been associated with low-density lipoprotein cholesterol (LDL-C) plasma levels^30^, associated with lower longevity ^31^ and with cognitive decline^32^. rs1160985 has been related to increased risk of LOAD in a Chinese population ^33 34^, but to be protective against AD in other ethnic cohorts^35 36^. Located in the APOE promoter region, the rs405509 minor allele in both copies has been described to alter APOE gene expression^37 38^ and to act as effect modifier to rs429358 in previous AD studies ^39 40 41^.

Intriguingly, the SNVs reaching the highest FI, rs1160985 and rs405509, had relatively low AF differences between LOAD and controls compared with the other prioritized SNVs. Also, the two most well-characterized LOAD genetic determinants in the literature to date and the ones with higher AF differences between both conditions, rs429358^27^ and rs7412^28^, were not the ones with the highest FI scores. These results suggest that tree-based ML methods are capable of prioritizing variants not only based on the individual enrichment of each SNV in the different classes, but also considering interactions between groups of SNVs.

When looking at the correctly classified genomic profiles, most of the TP were characterized to have rs429358, rs4420638 and rs769449 co-occurring in either one or two alleles, without the presence of rs405509 in two alleles and the absence of rs7412 (interaction 2). Contrarily, profiles with rs429358, rs4420638 and rs769449 in one allele co-occurring with rs405509 in two alleles were present in TN. In this sense, rs405509 seems to act as an effect modifier over the three predisposing variants. In addition, rs1160985 and rs405509 in both alleles were either predisposing to AD when co-occurring in TP (interaction 1), or protecting against AD when mutually exclusive in TN. Lastly, rs157580 and rs405697 were present in a small number of LOAD cases, comprising a third group of TP in GB. These two SNVs in TOMM40 were reported in other works to be related with lower longevity in Chinese population^42^ and related to AD independently of variants in the APOE locus in Japanese population^36^. When testing the statistical significance of the pairwise interactions in the set of 6SNVs prioritized by the three methods, results corroborated what we observed in the genomic profiles. The strongest interaction was present between rs405509 and rs1160985 followed by the interactions between rs405509 and the other SNVs located in APOE.

LOAD individuals with interaction 1 and interaction 2 were associated with specific comorbidities and an enrichment of diseases of the circulatory system was observed in both groups. Using GTEx data we show that the 6 prioritized SNVs are eQTLs of APOE or APOC1, but not in brain tissues. Conversely, rs1160985, rs405509, rs769449, rs4420638 are sQTL of TOMM40, and rs429358 is sQTL of TOMM40 and APOE in brain tissues. In this respect, some studies previously suggested the existence of a complex transcriptional regulatory hub in the region where the prioritized SNVs are located ^11 43 44^.

We cannot explain why the tree-based ML algorithms prioritized SNVs in the same chromosome19 region, ignoring other well reported AD genetic determinants such as rs744373 in BIN1^45^, rs3818361 in CR1^46^, or rs11136000 in CLU^47^. A possible reason could be that ML methods such as GB, ET and RF are efficient at classifying the major genetic profiles defined by a set of interactions between SNVs, ignoring other minor profiles caused by a single SNV.

As in other studies^17^, we found that tree-based ML methods are able to add an important layer of information to the disease related variants obtained with other population genomic approaches such as GWAS. In this regard, the combination of both methodologies could advance the discovery of new genomic profiles predisposing to the disease and the biological pathways involved. Furthermore, the validation of the genomic profiles could improve the clinical characterization of patients in the future. Nevertheless, the possibility of using individualized genomic information to stratify the population with the risk of developing a certain disease, especially if a cure is not yet available, is always controversial. With the balance of benefits and costs in mind, genetic tests could further the health care system implementing preventive measures in a healthy population with the risk of developing AD. Yet, an adequate regulation should be applied, considering topics such as personal data protection, privacy, and informed consent^48^.

## Data Availability

The data are available upon request to the first and corresponding authors

