## Supplementary Materials for "Machine learning methods applied to genotyping data capture interactions between single nucleotide variants in late onset Alzheimer’s disease"

**Supplementary Figure 2.** 10-fold cross-validation on the training set split into train and validation set in the hyperparameter optimization step. AUC-ROC (**a**, **b**, **c)** and F score (**d**, **e**, **f)** were used to evaluate the performance of the models and select the best parameters. The reference predictors as SNVs located in genes from DisGeNet "curated gene disease associations" dataset but not reported in the "curated variant disease associations" dataset using the AD categories in *Supplementary Table 1, page 2* were used to control for the over-fitting and to measure the specificity of the AD predictors to classify LOAD and controls.

**Supplementary Figure 3.** In **a** AUC-ROC obtained during the evaluation on the testing set with the three ML models. In **b**, **c**, **d** FI score for the 145 SNVs in AD predictors obtained in GB, ET and RF respectively, and the cut-off used to prioritize the SNVs.

**Supplementary Figure 4**. Genomic profiles of correctly classified samples in ET defined by the 20 prioritized SNVs. Same structure as in Figure 2.

**Supplementary Figure 5**. Genomic profiles of correctly classified samples in RF defined by the 15 prioritized SNVs. Same structure as in Figure 2.

#### Supplementary Tables

**Supplementary Table 1. page 1)** ICD-10 codes used to select Alzheimer's disease cases in UK Biobank.; **page 2)** Disease IDs in MedGen concept ID format (column Disease ID) used to select Alzheimer's disease related variants in DisGeNet database; **page 3)** Parameters selected in the three ML methods. Hyper-parameter selection was performed on the training set through a 10-fold cross-validation. Parameters not listed here were used as default in Scikit-learn module; **page 4)**

Percentage of AD and controls with genomic profiles in clusters captured by GB models (Figure 2). The percentages are calculated over the total AD n = 618 and Controls n= 62392 without any missing value in the prioritized SNVs. The differences between AD and controls are measured with a Fisher test; **page 5)** Information on the eQTLs obtained from GTEx portal in the 6 SNVs commonly prioritized by the three ML models; **page 6)** Information on the sQTLs obtained from GTEx portal in the 6 SNVs commonly prioritized by the three ML models.

**Supplementary Table 2.** SNVs from DisGenet used as AD predictors in this study. We report chromosome (Chr), position (hg19 Postition), reference allele (Ref), alternative allele (Alt), genetic region (Region) and gene ID (Gene Symbol).

**Supplementary Table 3.** SNVs selected by GB, ET and RF models (total of 9, 20 and 15, respectively). We report SNV ID (SNV), gene ID (Gene), genetic region (Region), chromosome (Chr), position (hg19 Position), allele frequency in Alzheimer's Disease (AF AD) and allele frequency in control group (AF Control) in testing and training sets (AF test AD, AF test Control, AF train AD, AF train Control) and the corresponding statistical metrics (LOG2 AF AD/Cntrl, FI and Fisher p-value).

**Supplementary Table 4**. Table with the diseases and conditions enriched in LOAD individuals with interaction profile 1 (rs1160985 and rs405509 in both alleles), interaction profile 2 (rs405509 not present in both alleles, rs769449 or rs429358 or rs429358 or rs4420638 present at least in one allele, rs7412 absent) with respect controls, or both profiles (Interact1, Interact2 and Intersect respectively). Fisher test was applied to measure the significance of the enrichment in the different subgroups, and a pval < 0.001 was used as a cut-off. Diseases and conditions present in more than 10% of LOAD individuals in the two subgroups (Interact1 and Interact2) are prioritized. We report ICD-10 codes (ICD10 codes), definition of the ICD-10 codes (meaning), proportion of LOAD with interaction 1 or 2 having the disease/condition (Prop.in.AD.WIth.InteractX), proportion of LOAD without the interaction having the disease/condition(Prop.in.OtherAD), proportion of controls having the disease/condition (Prop.in.Controls), p-value measuring the differences in the presence of the disease/condition in AD with interact 1 or 2 compared with other AD (pVal.ADinteractX.vs.OtherAD), p-value measuring the differences in the presence of the disease/condition in AD with interact 1 or 2 compared with controls (pVal.ADinteractX.vs.cntrl).

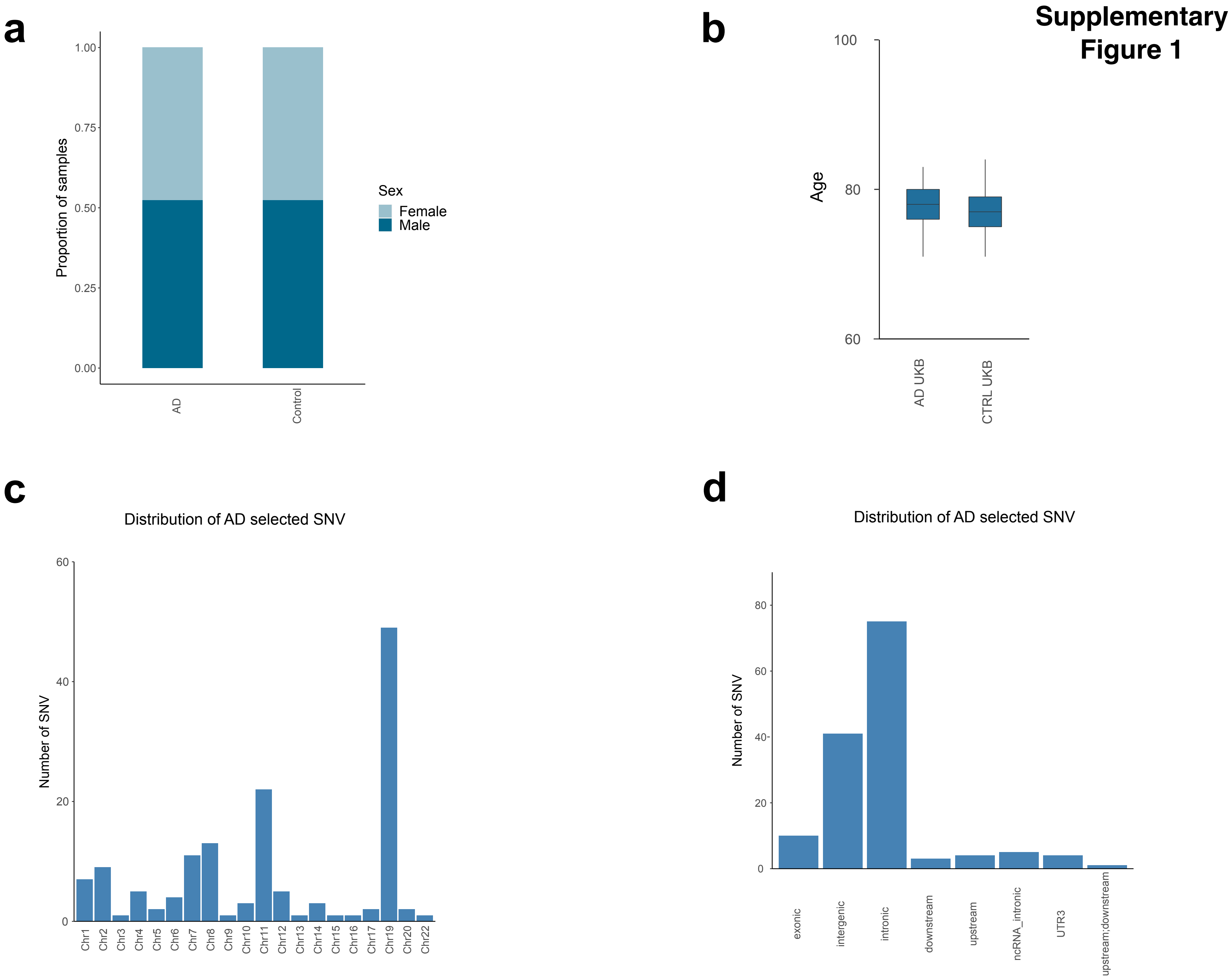
Supplementary Figure 1
a
Proportion of samples
1.00
0.75
0.50
0.25
0.00
AD
Control
Sex
Female
Male
b
Age
100
80
60
AD UKB
CTRL UKB
c
Distribution of AD selected SNV
Number of SNV
60
40
20
0
Chr1
Chr2
Chr3
Chr4
Chr5
Chr6
Chr7
Chr8
Chr9
Chr10
Chr11
Chr12
Chr13
Chr14
Chr15
Chr16
Chr17
Chr19
Chr20
Chr22
d
Distribution of AD selected SNV
Number of SNV
80
60
40
20
0
exonic
intergenic
intronic
downstream
upstream
ncRNA_intronic
UTR3
upstream;downstream

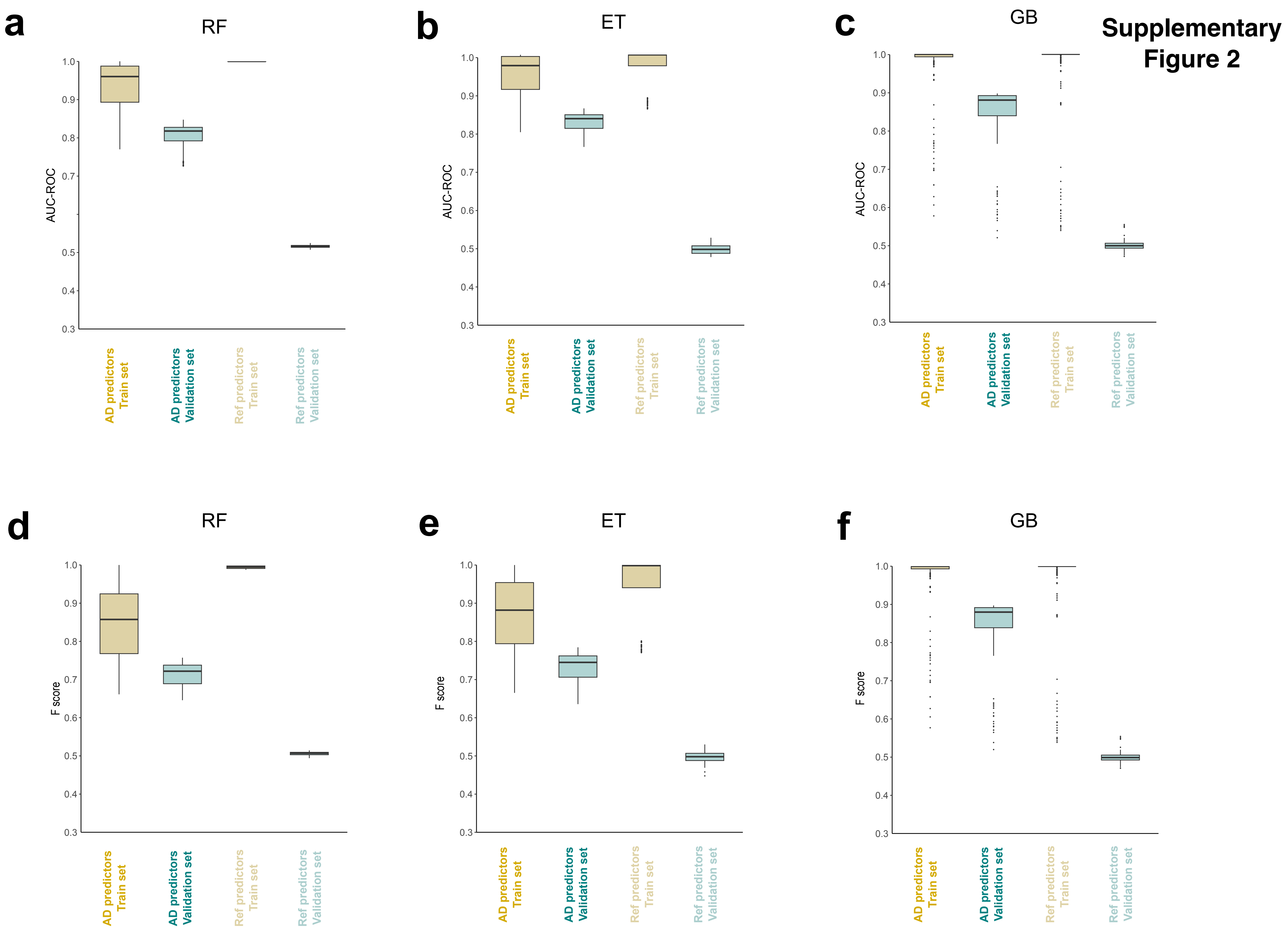

Supplementary Figure 2
a
RF
AUC-ROC
b
ET
AUC-ROC
c
GB
AUC-ROC
d
RF
F score
e
ET
F score
f
GB
F score
AD predictors Train set
AD predictors Validation set
Ref predictors Train set
Ref predictors Validation set

**Supplementary Figure 3**

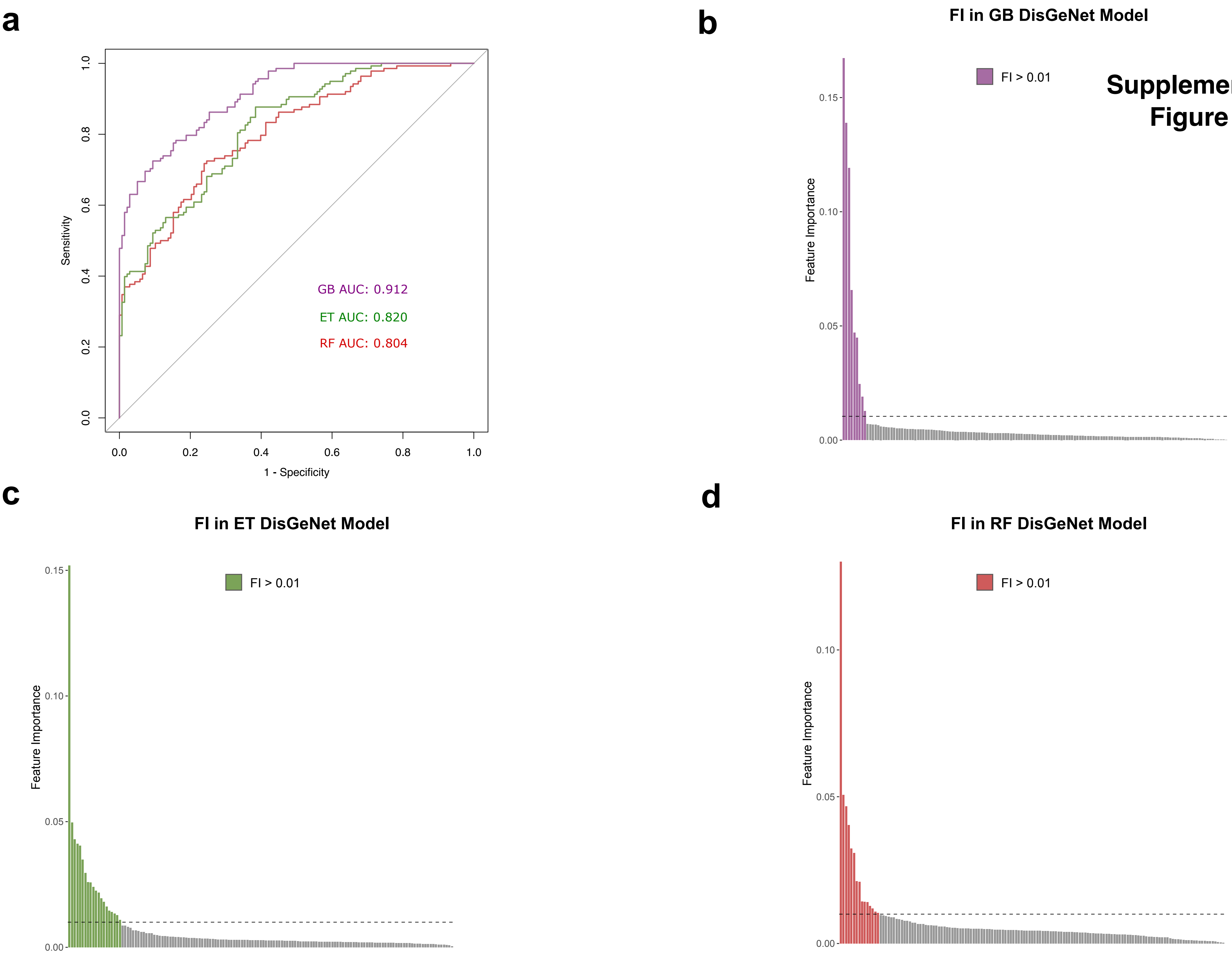

### Supplementary Figure 4

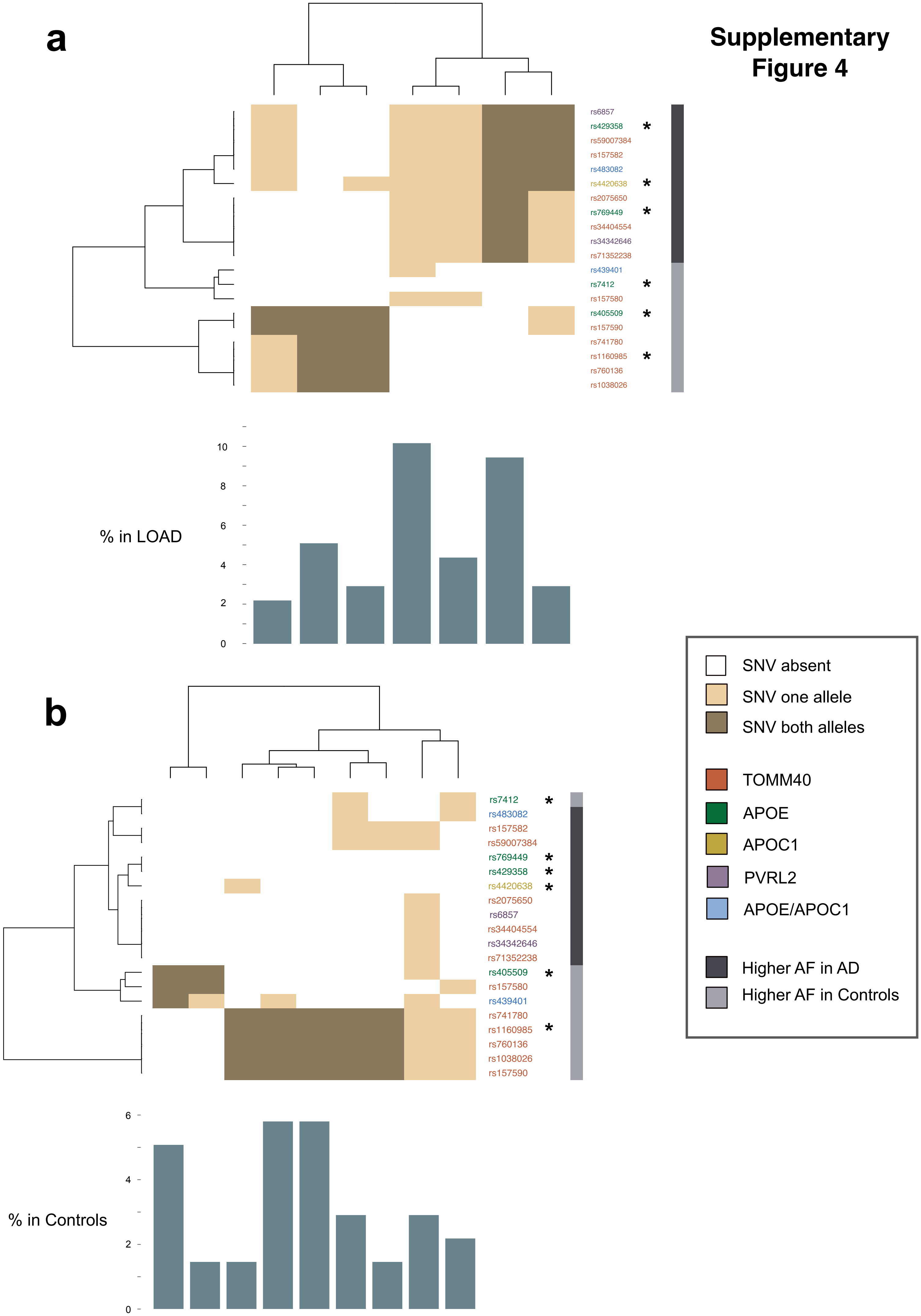

#### Supplementary Figure 5

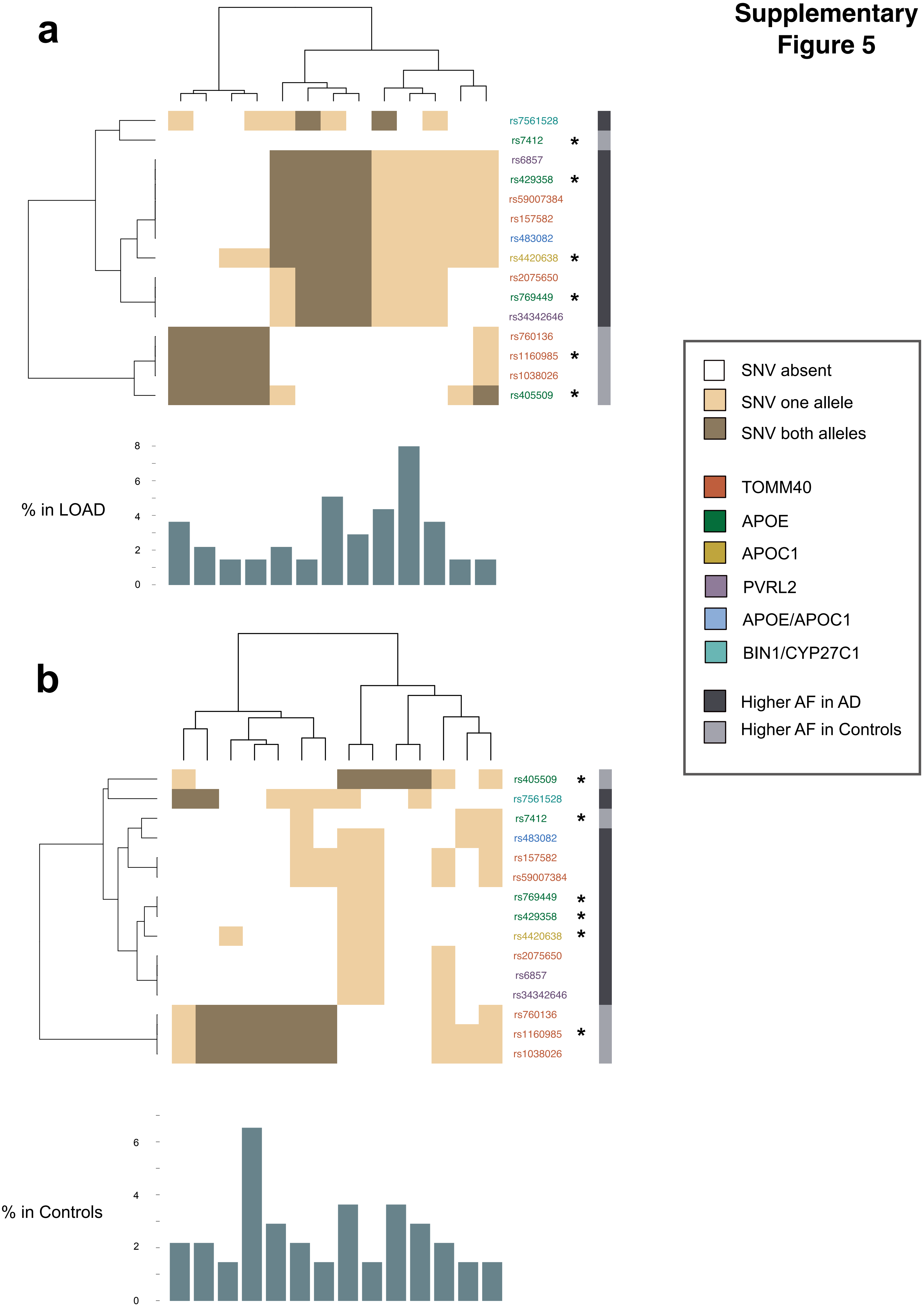

**Supplementary Table 1**

| ICD10 Coding | Meaning |
|---|---|
| F00 | F00 Dementia in Alzheimer's disease |
| F001 | F00.1 Dementia in Alzheimer's disease with late onset |
| F002 | F00.2 Dementia in Alzheimer's disease, atypical or mixed type |
| F009 | F00.9 Dementia in Alzheimer's disease, unspecified |
| G30 | G30 Alzheimer's disease |
| G301 | G30.1 Alzheimer's disease with late onset |
| G308 | G30.8 Other Alzheimer's disease |
| G309 | G30.9 Alzheimer's disease, unspecified |

ICD-10 codes used to select Alzheimer’s disease cases in UK Biobank

#### Supplementary Table 1

| Disease ID | Disease Name |
|---|---|
| C0002395 | Alzheimer's Disease |
| C1851958 | Lewy Body Variant of Alzheimer Disease |
| C1843013 | Alzheimer disease, familial, type 3 |
| C1843014 | Alzheimer Disease, Familial, 3, with Spastic Paraparesis and Unusual Plaques |
| C1843015 | Alzheimer Disease, Familial, 3, with Spastic Paraparesis and Apraxia |
| C0276496 | Familial Alzheimer Disease (FAD) |
| C0494463 | Alzheimer Disease, Late Onset |
| C0750900 | Alzheimer's Disease, Focal Onset |
| C1837149 | Alzheimer Disease 9 |

Disease IDs in MedGen concept ID format (column Disease ID) used to select Alzheimer's disease related variants in DisGeNet database.

**Supplementary Table 1**

| ML method | Parameters |
|---|---|
| Gradient Boosted Decision Trees (GB) | • n_estimators = 60<br>• min_samples_split = 5<br>• min_samples_leaf = 2<br>• max_depth = 7 |
| Extremely Randomized Trees (ET) | • n_estimators = 100<br>• min_samples_split = 8<br>• min_samples_leaf = 1<br>• max_depth = 7 |
| Random Forest (RF) | • n_estimators = 100<br>• learning_rate = 0.0001<br>• subsample = 0.7<br>• max_depth = 9<br>• loss = 'deviance' |

Parameters selected in the three ML methods. Hyper-parameter selection was performed on the training set with a 10-fold cross-validation. Parameters not listed here were used as default in Scikit-learn module.

**Supplementary Table 1**

| | | % LOAD | % Cntrl | Fisher pval |
|---|---|---|---|---|
| TP | C1 | 27.83 | 1.21 | 2.69E-168 |
| | C2 | 3.24 | 0.00 | 1.04E-39 |
| | C3 | 12.62 | 0.00 | 1.44E-159 |
| TN | C1 | 0.49 | 24.42 | 9.97E-52 |
| | C2 | 0.00 | 17.50 | 3.60E-69 |

Percentage of AD and Controls with genomic profiles in clusters captured by GB models (Figure 2). The percentages are calculated over the total AD n = 618 and Controls n= 62392 without any missing value in the prioritized SNVs. The differences between AD and controls are measured with a Fisher test.

**xd**

| Genomic loci | | | Changes in gene expression | |
|---|---|---|---|---|
| **SNV** | **Region** | **Gene Symbol** | **APOE** | **APOC1** |
| **rs1160985** | **intronic** | **TOMM40** | • Heart - Atrial Appendage: pVal1.3e-05<br>• Skin - Sun Exposed (Lower leg): pVal4e-05 | NA |
| **rs405509** | **upstream** | **APOE** | • Skin - Sun Exposed (Lower leg): pVal1.7e-08<br>• Skin - Not Sun Exposed (Suprapubic): pVal1.5e-07 | NA |
| **rs429358** | **exonic** | **APOE** | NA | • Esophagus – Mucosa: pVal2.3e-05<br>• Adrenal Gland: pVal4.8e-05 |
| **rs7412** | **exonic** | **APOE** | • Skin - Not Sun Exposed (Suprapubic): pVal1.2e-07 | NA |
| **rs769449** | **intronic** | **APOE** | NA | • Esophagus – Mucosa: pVal6.2e-05 |
| **rs4420638** | **downstream** | **APOC1** | • Skin - Not Sun Expose (Suprapubic): pVal2.4e-05<br>• Skin - Sun Exposed (Lower leg): pVal3.8e-05 | • Adrenal Gland: pVal1.2e-05<br>• Esophagus – Mucosa: pVal5.9e-05<br>• Skin - Sun Exposed (Lower leg): pVal6.9e-05 |

Information on the eQTLs obtained from GTEx portal in the 6 SNVs commonly prioritized by the three ML models.

#### Supplementary Table 1

| Genomic loci | | | Changes in splicing | |
|---|---|---|---|---|
| SNV | Region | Gene Symbol | TOMM40 | APOE |
| **rs1160985** | **intronic** | **TOMM40** | • Brain - Cerebellar Hemisphere: pVal1.3e-05 | NA |
| **rs405509** | **upstream** | **APOE** | • Brain - Cerebellar Hemisphere: pVal5.8e-06 | • Lung: pVal5.3e-06 |
| **rs429358** | **exonic** | **APOE** | • Brain - Cerebellar Hemisphere: pVal3.6e-11<br>• Brain – Cerebellum: pVal3e-06 | • Brain - Caudate (basal ganglia): pVal2e-06 |
| **rs7412** | **exonic** | **APOE** | NA | NA |
| **rs769449** | **intronic** | **APOE** | • Brain - Cerebellar Hemisphere: pVal4e-10<br>• Brain – Cerebellum: pVal1.5e-05 | NA |
| **rs4420638** | **downstream** | **APOC1** | • Brain - Cerebellar Hemisphere: pVal9.5e-07 | NA |

Information on the sQTLs obtained from GTEx portal in the 6 SNVs commonly prioritized by the three ML models.

### Supplementary Table 2

| dbSNP ID | Chr | hg19 Postition | Ref | Alt | Region | Gene Symbol | DisGeNet |
|---|---|---|---|---|---|---|---|
| rs1038026 | 19 | 45405062 | A | G | intronic | TOMM40 | YES |
| rs10405693 | 19 | 45326664 | C | T | intergenic | BCAM;PVRL2 | YES |
| rs10415983 | 19 | 45711598 | C | T | intergenic | BLOC1S3;EXOC3L2 | YES |
| rs10498633 | 14 | 92926952 | G | T | intronic | SLC24A4 | YES |
| rs10792832 | 11 | 85867875 | G | A | intergenic | PICALM;EED | YES |
| rs10850408 | 12 | 115380393 | C | T | intergenic | TBX3;MED13L | YES |
| rs10948363 | 6 | 47487762 | A | G | intronic | CD2AP | YES |
| rs11136000 | 8 | 27464519 | C | T | intronic | CLU | YES |
| rs11257238 | 10 | 11717397 | T | C | intergenic | USP6NL;ECHDC3 | YES |
| rs115786578 | 20 | 32737601 | G | A | intergenic | EIF2S2;ASIP | YES |
| rs1160985 | 19 | 45403412 | C | T | intronic | TOMM40 | YES |
| rs11610206 | 12 | 47639526 | T | C | intergenic | PCED1B;LOC105369747 | YES |
| rs11667640 | 19 | 45379791 | C | T | intronic | PVRL2 | YES |
| rs11669338 | 19 | 45382984 | T | G | intronic | PVRL2 | YES |
| rs11673139 | 19 | 45383037 | A | T | intronic | PVRL2 | YES |
| rs116932763 | 7 | 149549009 | G | A | intronic | ZNF862 | YES |
| rs11767557 | 7 | 143109139 | T | C | ncRNA_intronic | EPHA1-AS1 | YES |
| rs11770757 | 7 | 133747946 | G | A | intronic | EXOC4 | YES |
| rs11771145 | 7 | 143110762 | G | A | ncRNA_intronic | EPHA1-AS1 | YES |
| rs117964204 | 17 | 48692082 | C | T | intronic | CACNA1G | YES |
| rs117983694 | 11 | 85396880 | A | C | exonic | CCDC89 | YES |
| rs118071777 | 7 | 88244249 | G | A | intergenic | STEAP4;ZNF804B | YES |
| rs118152978 | 8 | 127353538 | A | G | intergenic | LOC101927657;FAM84B | YES |
| rs11824773 | 11 | 60076940 | G | C | downstream | MS4A4A | YES |
| rs12041233 | 1 | 37752707 | G | A | intergenic | MIR4255;LINC01137 | YES |
| rs12334143 | 7 | 85183016 | T | C | intergenic | LINC00972;GRM3 | YES |
| rs12361953 | 11 | 24611130 | C | G | intronic | LUZP2 | YES |
| rs12610605 | 19 | 45370838 | G | A | intronic | PVRL2 | YES |
| rs12787412 | 11 | 85703093 | C | T | intronic | PICALM | YES |
| rs12805520 | 11 | 85630411 | C | T | exonic | CCDC83 | YES |
| rs12989701 | 2 | 127887985 | C | A | intergenic | BIN1;CYP27C1 | YES |
| rs1385600 | 11 | 77936166 | A | G | exonic | GAB2 | YES |
| rs1466662 | 4 | 155347393 | A | T | intronic | DCHS2 | YES |
| rs1476679 | 7 | 100004446 | T | C | intronic | ZCWPW1 | YES |
| rs1531517 | 19 | 45242173 | G | A | intergenic | CEACAM16;BCL3 | YES |
| rs1532278 | 8 | 27466315 | C | T | intronic | CLU | YES |
| rs1562990 | 11 | 60023087 | A | C | intergenic | MS4A4E;MS4A4A | YES |
| rs157580 | 19 | 45395266 | A | G | intronic | TOMM40 | YES |
| rs157582 | 19 | 45396219 | C | T | intronic | TOMM40 | YES |
| rs157590 | 19 | 45398716 | A | C | intronic | TOMM40 | YES |

### Supplementary Table 2

| | | | | | | | |
|---|---|---|---|---|---|---|---|
| rs1582763 | 11 | 60021948 | G | A | intergenic | MS4A4E;MS4A4A | YES |
| rs16979595 | 19 | 45477381 | G | A | intronic | CLPTM1 | YES |
| rs17125944 | 14 | 53400629 | T | C | intronic | FERMT2 | YES |
| rs17817600 | 11 | 85677471 | A | G | intronic | PICALM | YES |
| rs1800795 | 7 | 22766645 | G | C | ncRNA_intronic | LOC541472 | YES |
| rs190982 | 5 | 88223420 | A | G | ncRNA_intronic | MEF2C-AS1 | YES |
| rs1925690 | 6 | 87867063 | C | T | intronic | ZNF292 | YES |
| rs2043948 | 14 | 75073048 | C | T | intronic | LTBP2 | YES |
| rs2075650 | 19 | 45395619 | A | G | intronic | TOMM40 | YES |
| rs2228467 | 3 | 42906116 | T | C | exonic | ACKR2 | YES |
| rs2279590 | 8 | 27456253 | C | T | intronic | CLU | YES |
| rs2373115 | 11 | 78091150 | C | A | intronic | GAB2 | YES |
| rs2421847 | 1 | 171557600 | A | G | exonic | PRRC2C | YES |
| rs2718058 | 7 | 37841534 | A | G | intergenic | GPR141;NME8 | YES |
| rs283813 | 19 | 45389174 | T | A | intronic | PVRL2 | YES |
| rs28834970 | 8 | 27195121 | T | C | intronic | PTK2B | YES |
| rs2965101 | 19 | 45237812 | T | C | intergenic | CEACAM16;BCL3 | YES |
| rs2965109 | 19 | 45225345 | C | T | intergenic | CEACAM16;BCL3 | YES |
| rs34342646 | 19 | 45388130 | G | A | intronic | PVRL2 | YES |
| rs34404554 | 19 | 45395909 | C | G | intronic | TOMM40 | YES |
| rs35349669 | 2 | 234068476 | C | T | intronic | INPP5D | YES |
| rs3745150 | 19 | 45385759 | G | C | intronic | PVRL2 | YES |
| rs3748140 | 8 | 8999019 | C | T | exonic | PPP1R3B | YES |
| rs3752246 | 19 | 1056492 | C | G | exonic | ABCA7 | YES |
| rs3760627 | 19 | 45457180 | T | C | upstream | CLPTM1 | YES |
| rs3763849 | 11 | 60193191 | C | T | intergenic | MS4A14;MS4A5 | YES |
| rs3764650 | 19 | 1046520 | T | G | intronic | ABCA7 | YES |
| rs3781834 | 11 | 121445940 | A | G | intronic | SORL1 | YES |
| rs3794318 | 12 | 8758543 | A | G | intronic | AICDA | YES |
| rs3818361 | 1 | 207784968 | G | A | intronic | CR1 | YES |
| rs3851179 | 11 | 85868640 | C | T | intergenic | PICALM;EED | YES |
| rs3865444 | 19 | 51727962 | C | A | upstream | CD33 | YES |
| rs387976 | 19 | 45379060 | A | C | intronic | PVRL2 | YES |
| rs3931397 | 4 | 149079497 | G | T | intronic | NR3C2 | YES |
| rs4038129 | 2 | 17774746 | A | G | intronic | VSNL1 | YES |
| rs4038131 | 2 | 17775032 | A | G | intronic | VSNL1 | YES |
| rs405509 | 19 | 45408836 | G | T | upstream | APOE | YES |
| rs405697 | 19 | 45404691 | G | A | intronic | TOMM40 | YES |
| rs4147929 | 19 | 1063443 | G | A | intronic | ABCA7 | YES |
| rs416041 | 19 | 45370854 | A | G | intronic | PVRL2 | YES |
| rs429358 | 19 | 45411941 | T | C | exonic | APOE | YES |
| rs439401 | 19 | 45414451 | C | T | intergenic | APOE;APOC1 | YES |

### Supplementary Table 2

| | | | | | | | |
|---|---|---|---|---|---|---|---|
| rs440277 | 19 | 45361224 | G | A | intronic | PVRL2 | YES |
| rs4420638 | 19 | 45422946 | A | G | downstream | APOC1 | YES |
| rs4557697 | 8 | 63843631 | A | G | intronic | NKAIN3 | YES |
| rs4663098 | 2 | 127873035 | C | T | intergenic | BIN1;CYP27C1 | YES |
| rs4676049 | 2 | 109635257 | C | T | intergenic | EDAR;SH3RF3-AS1 | YES |
| rs471470 | 11 | 85831541 | A | C | intergenic | PICALM;EED | YES |
| rs4803770 | 19 | 45427353 | C | G | intergenic | APOC1;APOC1P1 | YES |
| rs483082 | 19 | 45416178 | G | T | intergenic | APOE;APOC1 | YES |
| rs4844610 | 1 | 207802552 | C | A | intronic | CR1 | YES |
| rs4938933 | 11 | 60034429 | T | C | intergenic | MS4A4E;MS4A4A | YES |
| rs519113 | 19 | 45376284 | C | G | intronic | PVRL2 | YES |
| rs519825 | 19 | 45366779 | T | C | intronic | PVRL2 | YES |
| rs536841 | 11 | 85787824 | T | C | intergenic | PICALM;EED | YES |
| rs541458 | 11 | 85788351 | T | C | intergenic | PICALM;EED | YES |
| rs56039743 | 1 | 26620806 | T | C | exonic | UBXN11 | YES |
| rs561655 | 11 | 85800279 | A | G | intergenic | PICALM;EED | YES |
| rs569214 | 8 | 27487790 | G | T | intergenic | CLU;SCARA3 | YES |
| rs59007384 | 19 | 45396665 | G | T | intronic | TOMM40 | YES |
| rs6088727 | 20 | 33713639 | A | G | intronic | EDEM2 | YES |
| rs610932 | 11 | 59939307 | G | T | UTR3 | MS4A6A | YES |
| rs611267 | 11 | 60005268 | G | A | intronic | MS4A4E | YES |
| rs61510607 | 7 | 155224715 | T | C | intergenic | LOC100286906;EN2 | YES |
| rs61812598 | 1 | 154420087 | G | A | intronic | IL6R | YES |
| rs624290 | 9 | 3928115 | T | C | intronic | GLIS3 | YES |
| rs639012 | 11 | 85681583 | G | A | intronic | PICALM | YES |
| rs6431223 | 2 | 127895487 | G | A | intergenic | BIN1;CYP27C1 | YES |
| rs6448799 | 4 | 11630049 | C | T | intergenic | HS3ST1;LINC02360 | YES |
| rs6701713 | 1 | 207786289 | G | A | intronic | CR1 | YES |
| rs6834555 | 4 | 10062326 | A | G | intergenic | SLC2A9;WDR1 | YES |
| rs6857 | 19 | 45392254 | C | T | UTR3 | PVRL2 | YES |
| rs6859 | 19 | 45382034 | G | A | UTR3 | PVRL2 | YES |
| rs690705 | 13 | 34654918 | A | G | intergenic | RFC3;LINC02343 | YES |
| rs6922617 | 6 | 41336101 | G | A | intergenic | NCR2;LINC01276 | YES |
| rs7009219 | 8 | 53214265 | C | T | intronic | ST18 | YES |
| rs7011581 | 8 | 9000465 | G | A | intronic | PPP1R3B | YES |
| rs7081208 | 10 | 13991865 | G | A | intronic | FRMD4A | YES |
| rs71352238 | 19 | 45394336 | T | C | upstream | TOMM40 | YES |
| rs714948 | 19 | 45165912 | C | A | UTR3 | PVR | YES |
| rs7225151 | 17 | 5137047 | G | A | ncRNA_intronic | LOC100130950 | YES |
| rs7259620 | 19 | 45407788 | G | A | downstream | TOMM40 | YES |
| rs7274581 | 20 | 55018260 | T | C | intronic | CASS4 | YES |

### Supplementary
### Table 2

| | | | | | | | |
|---|---|---|---|---|---|---|---|
| rs72807343 | 5 | 179238261 | C | T | intronic | SQSTM1 | YES |
| rs7295246 | 12 | 43967677 | T | G | intergenic | ADAMTS20;PUS7L | YES |
| rs7412 | 19 | 45412079 | C | T | exonic | APOE | YES |
| rs741780 | 19 | 45404431 | T | C | intronic | TOMM40 | YES |
| rs744373 | 2 | 127894615 | A | G | intergenic | BIN1;CYP27C1 | YES |
| rs74495807 | 7 | 34088535 | C | T | intronic | BMPER | YES |
| rs749005 | 6 | 6283666 | G | T | intronic | F13A1 | YES |
| rs753129 | 4 | 56668431 | A | G | intergenic | NMU;LOC644145 | YES |
| rs7561528 | 2 | 127889637 | G | A | intergenic | BIN1;CYP27C1 | YES |
| rs75617873 | 22 | 44526105 | A | C | intronic | PARVB | YES |
| rs760136 | 19 | 45403858 | A | G | intronic | TOMM40 | YES |
| rs769449 | 19 | 45410002 | G | A | intronic | APOE | YES |
| rs7818382 | 8 | 96054000 | C | T | intronic | NDUFAF6 | YES |
| rs7920721 | 10 | 11720308 | A | G | intergenic | USP6NL;ECHDC3 | YES |
| rs7933202 | 11 | 59936926 | A | C | intergenic | MS4A2;MS4A6A | YES |
| rs79335261 | 12 | 56497641 | G | T | upstream;downstream | PA2G4;ERBB3 | YES |
| rs8035452 | 15 | 51040798 | T | C | intronic | SPPL2A | YES |
| rs8105340 | 19 | 45367777 | T | C | intronic | PVRL2 | YES |
| rs8106922 | 19 | 45401666 | A | G | intronic | TOMM40 | YES |
| rs9302457 | 16 | 11059837 | G | A | intronic | CLEC16A | YES |
| rs9331888 | 8 | 27468862 | C | G | intronic | CLU | YES |
| rs9331896 | 8 | 27467686 | T | C | intronic | CLU | YES |

### Supplementary Table 3

| SNV | Gene | Region | Chr | hg19 Postition | AF AD | AF Control | AF test AD | AF test Control | AF train AD | AF train Control | LOG2 AF AD/Cntrl | FI | Fisher p.val |
|---|---|---|---|---|---|---|---|---|---|---|---|---|---|
| **rs429358** | APOE | exonic | 19 | 45411941 | 0,403 | 0,156 | 0,400 | 0,179 | 0,404 | 0,150 | 1,373 | 0,066 | 3,72E-44 |
| **rs769449** | APOE | intronic | 19 | 45410002 | 0,320 | 0,127 | 0,338 | 0,145 | 0,315 | 0,123 | 1,329 | 0,119 | 9,95E-37 |
| **rs4420638** | APOC1 | downstream | 19 | 45422946 | 0,417 | 0,189 | 0,438 | 0,201 | 0,412 | 0,186 | 1,144 | 0,045 | 4,56E-42 |
| **rs405509** | APOE | upstream | 19 | 45408836 | 0,411 | 0,470 | 0,380 | 0,471 | 0,418 | 0,470 | -0,194 | 0,139 | 0,00140019 |
| rs157590 | TOMM40 | intronic | 19 | 45398716 | 0,400 | 0,480 | 0,376 | 0,516 | 0,405 | 0,472 | -0,264 | 0,019 | 3,72E-05 |
| rs157580 | TOMM40 | intronic | 19 | 45395266 | 0,282 | 0,378 | 0,268 | 0,326 | 0,285 | 0,390 | -0,425 | 0,013 | 3,18E-08 |
| rs405697 | TOMM40 | intronic | 19 | 45404691 | 0,199 | 0,268 | 0,185 | 0,246 | 0,202 | 0,273 | -0,428 | 0,047 | 3,27E-05 |
| **rs1160985** | TOMM40 | intronic | 19 | 45403412 | 0,322 | 0,458 | 0,310 | 0,478 | 0,325 | 0,453 | -0,507 | 0,167 | 5,04E-14 |
| **rs7412** | APOE | exonic | 19 | 45412079 | 0,033 | 0,087 | 0,036 | 0,073 | 0,032 | 0,090 | -1,385 | 0,024 | 5,14E-09 |

### Supplementary Table 3

| SNV | Gene | Region | Chr | hg19 Position | AF AD | AF Control | AF test AD | AF test Control | AF train AD | AF train Control | LOG2 AF AD/Cntrl | FI | Fisher p.val |
|---|---|---|---|---|---|---|---|---|---|---|---|---|---|
| **rs429358** | APOE | exonic | 19 | 45411941 | 0,403 | 0,156 | 0,400 | 0,179 | 0,404 | 0,150 | 1,373 | 0,041 | 3,72E-44 |
| **rs769449** | APOE | intronic | 19 | 45410002 | 0,320 | 0,127 | 0,338 | 0,145 | 0,315 | 0,123 | 1,329 | 0,043 | 9,95E-37 |
| rs6857 | PVRL2 | UTR3 | 19 | 45392254 | 0,383 | 0,164 | 0,411 | 0,187 | 0,377 | 0,159 | 1,224 | 0,035 | 5,11E-37 |
| rs2075650 | TOMM40 | intronic | 19 | 45395619 | 0,320 | 0,139 | 0,355 | 0,163 | 0,312 | 0,134 | 1,203 | 0,026 | 5,41E-32 |
| rs34404554 | TOMM40 | intronic | 19 | 45395909 | 0,326 | 0,144 | 0,368 | 0,165 | 0,316 | 0,139 | 1,180 | 0,020 | 1,05E-28 |
| rs34342646 | PVRL2 | intronic | 19 | 45388130 | 0,327 | 0,145 | 0,375 | 0,165 | 0,316 | 0,141 | 1,170 | 0,023 | 2,01E-28 |
| rs71352238 | TOMM40 | upstream | 19 | 45394336 | 0,322 | 0,145 | 0,368 | 0,169 | 0,312 | 0,139 | 1,153 | 0,024 | 2,66E-27 |
| **rs4420638** | APOC1 | downstream | 19 | 45422946 | 0,417 | 0,189 | 0,438 | 0,201 | 0,412 | 0,186 | 1,144 | 0,050 | 4,56E-42 |
| rs59007384 | TOMM40 | intronic | 19 | 45396665 | 0,408 | 0,213 | 0,432 | 0,246 | 0,403 | 0,205 | 0,940 | 0,041 | 1,49E-27 |
| rs157582 | TOMM40 | intronic | 19 | 45396219 | 0,419 | 0,223 | 0,440 | 0,252 | 0,414 | 0,216 | 0,907 | 0,022 | 4,13E-27 |
| rs483082 | APOE;APOC1 | intergenic | 19 | 45416178 | 0,432 | 0,246 | 0,447 | 0,246 | 0,429 | 0,246 | 0,813 | 0,030 | 5,29E-24 |
| **rs405509** | APOE | upstream | 19 | 45408836 | 0,411 | 0,470 | 0,380 | 0,471 | 0,418 | 0,470 | -0,194 | 0,152 | 0,00140019 |
| rs157590 | TOMM40 | intronic | 19 | 45398716 | 0,400 | 0,480 | 0,376 | 0,516 | 0,405 | 0,472 | -0,264 | 0,014 | 3,72E-05 |
| rs439401 | APOE;APOC1 | intergenic | 19 | 45414451 | 0,261 | 0,343 | 0,268 | 0,359 | 0,259 | 0,339 | -0,396 | 0,013 | 1,39E-06 |
| rs157580 | TOMM40 | intronic | 19 | 45395266 | 0,282 | 0,378 | 0,268 | 0,326 | 0,285 | 0,390 | -0,425 | 0,016 | 3,18E-08 |
| rs1038026 | TOMM40 | intronic | 19 | 45405062 | 0,322 | 0,449 | 0,308 | 0,476 | 0,325 | 0,442 | -0,480 | 0,011 | 2,19E-11 |
| rs741780 | TOMM40 | intronic | 19 | 45404431 | 0,321 | 0,448 | 0,308 | 0,476 | 0,324 | 0,442 | -0,481 | 0,013 | 2,15E-11 |
| rs760136 | TOMM40 | intronic | 19 | 45403858 | 0,321 | 0,448 | 0,308 | 0,476 | 0,324 | 0,442 | -0,481 | 0,018 | 2,15E-11 |
| **rs1160985** | TOMM40 | intronic | 19 | 45403412 | 0,322 | 0,458 | 0,310 | 0,478 | 0,325 | 0,453 | -0,507 | 0,026 | 5,04E-14 |
| **rs7412** | APOE | exonic | 19 | 45412079 | 0,033 | 0,087 | 0,036 | 0,073 | 0,032 | 0,090 | -1,385 | 0,015 | 5,14E-09 |

### Supplementary Table 3

| SNV | Gene | Region | Chr | hg19 Position | AF AD | AF Control | AF test AD | AF test Control | AF train AD | AF train Control | LOG2 AF AD/Cntrl | FI | Fisher p.val |
|---|---|---|---|---|---|---|---|---|---|---|---|---|---|
| **rs429358** | APOE | exonic | 19 | 45411941 | 0,403 | 0,156 | 0,400 | 0,179 | 0,404 | 0,150 | 1,373 | 0,047 | 3,72E-44 |
| **rs769449** | APOE | intronic | 19 | 45410002 | 0,320 | 0,127 | 0,338 | 0,145 | 0,315 | 0,123 | 1,329 | 0,040 | 9,95E-37 |
| rs6857 | PVRL2 | UTR3 | 19 | 45392254 | 0,383 | 0,164 | 0,411 | 0,187 | 0,377 | 0,159 | 1,224 | 0,032 | 5,11E-37 |
| rs2075650 | TOMM40 | intronic | 19 | 45395619 | 0,320 | 0,139 | 0,355 | 0,163 | 0,312 | 0,134 | 1,203 | 0,021 | 5,41E-32 |
| rs34342646 | PVRL2 | intronic | 19 | 45388130 | 0,327 | 0,145 | 0,375 | 0,165 | 0,316 | 0,141 | 1,170 | 0,014 | 2,01E-28 |
| **rs4420638** | APOC1 | downstream | 19 | 45422946 | 0,417 | 0,189 | 0,438 | 0,201 | 0,412 | 0,186 | 1,144 | 0,051 | 4,56E-42 |
| rs59007384 | TOMM40 | intronic | 19 | 45396665 | 0,408 | 0,213 | 0,432 | 0,246 | 0,403 | 0,205 | 0,940 | 0,021 | 1,49E-27 |
| rs157582 | TOMM40 | intronic | 19 | 45396219 | 0,419 | 0,223 | 0,440 | 0,252 | 0,414 | 0,216 | 0,907 | 0,031 | 4,13E-27 |
| rs483082 | APOE;APOC1 | intergenic | 19 | 45416178 | 0,432 | 0,246 | 0,447 | 0,246 | 0,429 | 0,246 | 0,813 | 0,014 | 5,29E-24 |
| rs7561528 | BIN1;CYP27C 1 | intergenic | 2 | 127889637 | 0,375 | 0,305 | 0,326 | 0,304 | 0,387 | 0,306 | 0,298 | 0,010 | 6,24E-05 |
| **rs405509** | APOE | upstream | 19 | 45408836 | 0,411 | 0,470 | 0,380 | 0,471 | 0,418 | 0,470 | -0,194 | 0,130 | 0,00140019 |
| rs1038026 | TOMM40 | intronic | 19 | 45405062 | 0,322 | 0,449 | 0,308 | 0,476 | 0,325 | 0,442 | -0,480 | 0,011 | 2,19E-11 |
| rs760136 | TOMM40 | intronic | 19 | 45403858 | 0,321 | 0,448 | 0,308 | 0,476 | 0,324 | 0,442 | -0,481 | 0,012 | 2,15E-11 |
| **rs1160985** | TOMM40 | intronic | 19 | 45403412 | 0,322 | 0,458 | 0,310 | 0,478 | 0,325 | 0,453 | -0,507 | 0,013 | 5,04E-14 |
| **rs7412** | APOE | exonic | 19 | 45412079 | 0,033 | 0,087 | 0,036 | 0,073 | 0,032 | 0,090 | -1,385 | 0,014 | 5,14E-09 |

### Supplementary Table 4

| ICD10 codes | meaning | Prop.in.AD.WIth.Interact1 | Prop.in.OtherAD | Prop.in.Controls | pVal.ADinteract1.vs.OtherAD | pVal.ADinteract1.vs.cntrl | Prop.in.AD.WIth.Interact2 | Prop.in.OtherAD | Prop.in.Controls | pVal.ADinteract2.vs.OtherAD | pVal.ADinteract2.vs.cntrl |
|---|---|---|---|---|---|---|---|---|---|---|---|
| **M1399** | **M13.99 Arthritis, unspecified (Site unspecified)** | **0,10126582** | **0,02896341** | **0,01354335** | **0,0050824** | **1,29E-05** | **0,02432432** | **0,05928854** | **0,01367985** | **0,03321474** | **0,10820799** |
| **R31** | **R31 Unspecified haematuria** | **0,17721519** | **0,08993902** | **0,05725145** | **0,02588573** | **0,00014925** | **0,09189189** | **0,11857708** | **0,05757334** | **0,28572342** | **0,00955968** |
| **K297** | **K29.7 Gastritis, unspecified** | **0,15189873** | **0,07317073** | **0,04602331** | **0,02662433** | **0,000263** | **0,08108108** | **0,09486166** | **0,04563138** | **0,56437871** | **0,00255735** |
| Z538 | Z53.8 Procedure not carried out for other reasons | 0,15189873 | 0,07621951 | 0,04682628 | 0,03078886 | 0,00030754 | 0,08108108 | 0,1027668 | 0,0458227 | 0,39290936 | 0,00265519 |

### Supplementary Table 4

| ICD10 codes | meaning | Prop.in.AD.WIth.Interact1 | Prop.in.OtherAD | Prop.in.Controls | pVal.ADinteract1.vs.OtherAD | pVal.ADinteract1.vs.cntrl | Prop.in.AD.WIth.Interact2 | Prop.in.OtherAD | Prop.in.Controls | pVal.ADinteract2.vs.OtherAD | pVal.ADinteract2.vs.cntrl |
|---|---|---|---|---|---|---|---|---|---|---|---|
| **R634** | **R63.4 Abnormal weight loss** | **0,06329114** | **0,09756098** | **0,0175448** | **0,41595585** | **0,01291874** | **0,11351351** | **0,08300395** | **0,01725128** | **0,2265439** | **2,88E-21** |
| **J22** | **J22 Unspecified acute lower respiratory infection** | **0,07594937** | **0,13414634** | **0,02384808** | **0,15763059** | **0,01169761** | **0,11891892** | **0,13833992** | **0,02302296** | **0,54011422** | **2,16E-18** |
| I259 | **I25.9 Chronic ischaemic heart disease, unspecified** | 0,13924051 | 0,1722561 | 0,04900767 | 0,5274105 | 0,00163271 | 0,16486486 | 0,16205534 | 0,04684311 | 1 | 2,64E-17 |
| I252 | **I25.2 Old myocardial infarction** | 0,10126582 | 0,11280488 | 0,03341675 | 0,85208165 | 0,00496362 | 0,11351351 | 0,12252964 | 0,03217474 | 0,79995329 | 3,57E-12 |
| **E039** | **E03.9 Hypothyroidism, unspecified** | **0,05063291** | **0,10365854** | **0,04341367** | **0,16195281** | **0,77811276** | **0,10810811** | **0,09881423** | **0,04347895** | **0,78997587** | **2,46E-07** |
| I209 | I20.9 Angina pectoris, unspecified | 0,16455696 | 0,1554878 | 0,06276515 | 0,86969005 | 0,00124696 | 0,12972973 | 0,17786561 | 0,06098533 | 0,10925837 | 1,09E-06 |
| **Z921** | **Z92.1 Personal history of long-term (current) use of anticoagulants** | **0,07594937** | **0,13109756** | **0,0485928** | **0,20718146** | **0,28413829** | **0,10540541** | **0,13833992** | **0,04754464** | **0,25632513** | **6,54E-06** |
| **I251** | **I25.1 Atherosclerotic heart disease** | **0,10126582** | **0,12957317** | **0,08069804** | **0,59178464** | **0,53126082** | **0,14594595** | **0,11067194** | **0,07869898** | **0,22815554** | **1,63E-05** |
| I48 | I48 Atrial fibrillation and flutter | 0,11392405 | 0,1554878 | 0,0696305 | 0,40655177 | 0,12116152 | 0,12162162 | 0,17391304 | 0,06825574 | 0,08006736 | 0,00018284 |

Diseases highlighted with this color seem to be also enriched in Interact1, but are near the cut-off

### Supplementary Table 4

| ICD10 codes | meaning | Prop.in.AD.WIth.Interact1 | Prop.in.OtherAD | Prop.in.Controls | pVal.ADinteract1.vs.OtherAD | pVal.ADinteract1.vs.cntrl | Prop.in.AD.WIth.Interact2 | Prop.in.OtherAD | Prop.in.Controls | pVal.ADinteract2.vs.OtherAD | pVal.ADinteract2.vs.cntrl |
|---|---|---|---|---|---|---|---|---|---|---|---|
| **N390** | **N39.0 Urinary tract infection, site not specified** | **0,29113924** | **0,31097561** | **0,03680259** | **0,79707273** | **7,34E-15** | **0,33783784** | **0,30039526** | **0,03628827** | **0,33832014** | **5,07E-82** |
| R296 | R29.6 Tendency to fall, not elsewhere classified | 0,18987342 | 0,20426829 | 0,00517913 | 0,88244154 | 2,74E-19 | 0,18648649 | 0,21343874 | 0,00511798 | 0,41400935 | 1,56E-80 |
| R268 | R26.8 Other and unspecified abnormalities of gait and mobility | 0,20253165 | 0,14634146 | 0,00364011 | 0,18735378 | 2,65E-23 | 0,12972973 | 0,17786561 | 0,00357143 | 0,10925837 | 7,23E-56 |
| R410 | R41.0 Disorientation, unspecified | 0,18987342 | 0,16158537 | 0,00556723 | 0,52157409 | 7,74E-19 | 0,13513514 | 0,2055336 | 0,00550064 | 0,02095841 | 1,27E-50 |
| R32 | R32 Unspecified urinary incontinence | 0,15189873 | 0,11890244 | 0,0053531 | 0,36831863 | 2,44E-14 | 0,11621622 | 0,13833992 | 0,00530931 | 0,45991322 | 1,17E-41 |
| E86 | E86 Volume depletion | 0,16455696 | 0,10823171 | 0,0092341 | 0,13681907 | 5,98E-13 | 0,10540541 | 0,11857708 | 0,00900829 | 0,60585324 | 2,56E-28 |
| **I10** | **I10 Essential (primary) hypertension** | **0,50632911** | **0,57317073** | **0,30273142** | **0,28038566** | **0,00018845** | **0,57027027** | **0,55731225** | **0,29963329** | **0,80506976** | **7,60E-27** |
| R55 | R55 Syncope and collapse | 0,2278481 | 0,17987805 | 0,02914765 | 0,28678104 | 1,23E-11 | 0,15945946 | 0,21343874 | 0,02916135 | 0,09106474 | 1,11E-25 |
| **N179** | **N17.9 Acute renal failure, unspecified** | **0,25316456** | **0,16158537** | **0,02139903** | **0,05637453** | **3,58E-16** | **0,13513514** | **0,21343874** | **0,02064732** | **0,01179131** | **4,32E-25** |
| K590 | K59.0 Constipation | 0,21518987 | 0,16006098 | 0,03175729 | 0,20432625 | 4,34E-10 | 0,15135135 | 0,18181818 | 0,0309949 | 0,32280073 | 3,94E-22 |
| **Z867** | **Z86.7 Personal history of diseases of the circulatory system** | **0,25316456** | **0,21341463** | **0,05334368** | **0,47021142** | **4,46E-09** | **0,18918919** | **0,25296443** | **0,05288584** | **0,05990181** | **3,26E-20** |
| **E780** | **E78.0 Pure hypercholesterolaemia** | **0,3164557** | **0,28963415** | **0,12455335** | **0,60322357** | **6,56E-06** | **0,29459459** | **0,28458498** | **0,12241709** | **0,85743446** | **2,24E-18** |
| **D649** | **D64.9 Anaemia, unspecified** | **0,13924051** | **0,13719512** | **0,03713716** | **1** | **0,0001647** | **0,12972973** | **0,15019763** | **0,03679847** | **0,47979392** | **9,02E-14** |
| **Z864** | **Z86.4 Personal history of psychoactive substance abuse** | **0,30379747** | **0,26219512** | **0,10410449** | **0,4218806** | **9,61E-07** | **0,23243243** | **0,28458498** | **0,10165816** | **0,15965095** | **3,66E-13** |
| R074 | R07.4 Chest pain, unspecified | 0,17721519 | 0,17682927 | 0,06570935 | 1 | 0,00060184 | 0,17297297 | 0,18181818 | 0,06454082 | 0,83070929 | 9,40E-13 |
| E119 | E11.9 Without complications | 0,18987342 | 0,19054878 | 0,06746249 | 1 | 0,00023706 | 0,15405405 | 0,24901186 | 0,06519452 | 0,00373408 | 2,36E-09 |
| Z922 | Z92.2 Personal history of long-term (current) use of other medicaments | 0,20253165 | 0,16006098 | 0,06715469 | 0,33662654 | 6,34E-05 | 0,14054054 | 0,19367589 | 0,06619898 | 0,09650831 | 5,08E-07 |
